# µ-Opioid Modulation of Sensorimotor Functional Connectivity in Autism: Insights from a Pharmacological Neuroimaging Investigation using Tianeptine

**DOI:** 10.1101/2025.03.12.25323848

**Authors:** Mihail Dimitrov, Nichol M.L. Wong, Sydney Leaman, Lucas G. S. França, Ioannis Valasakis, Jason He, David J. Lythgoe, James L. Findon, Robert H. Wichers, Vladimira Stoencheva, Dene M. Robertson, Sarah Blainey, Glynis Ivin, Štefan Holiga, Mark D. Tricklebank, Dafnis Batalle, Declan G.M. Murphy, Gráinne M. McAlonan, Eileen Daly

## Abstract

Reproducible patterns of atypical functional connectivity of sensorimotor and higher-order networks have been previously identified in the autistic brain. However, the neurosignalling pathways underpinning these differences remain unclear. The µ-opioid system is involved in sensory processing as well as social and reward behaviours and has been implicated in autism, suggesting a potential role in shaping the autistic brain. Hence, we tested the hypothesis that there is atypical involvement of the µ-opioid system in these networks in autism. We used a placebo-controlled, double-blind, randomised, crossover study design to compare the effects of an acute dose of the µ-opioid receptor agonist tianeptine in autistic and non-autistic participants on functional connectivity (FC) of sensorimotor and frontoparietal networks. We found that tianeptine increased FC of a sensorimotor network previously characterised by atypically *low* FC in autism. The connectivity of the frontoparietal network was not significantly shifted. Our findings suggest that µ-opioid neurosignalling might contribute to functional brain differences in the sensorimotor network in autism. Given that sensorimotor system alterations are thought to be core to autism and contribute to other core autistic features, as well as adaptability and mental health, further research is warranted to explore the translational potential of µ-opioid modulation in autism.

## Introduction

Autism is a heterogeneous spectrum of neurodevelopmental conditions that is characterised by social communication differences, and repetitive and stereotyped behaviours, including sensory atypicalities^1^. It is present in up to 2.8% of the population^1–4^ and is associated with an increased likelihood of physical and mental health conditions (such as depression and anxiety), higher rates of unemployment, and lower overall wellbeing and quality of life^1^. Nevertheless, pharmacological intervention options for those who would like that choice are still largely unavailable^1, 5^ as are validated and meaningful stratification and candidate drug response biomarkers^6^. This might in part be due to our incomplete understanding of autism neurobiology.

Multiple prior hypotheses propose that the neurobiological underpinnings of autistic behaviours pivot around differences in brain connectivity. Recent large-scale resting-state functional Magnetic Resonance Imaging (rs-fMRI) studies have reported subtle but reproducible differences across different data sets in so-called resting-state functional connectivity (rsFC); with over-connectivity of higher-order networks and under-connectivity of sensorimotor networks^7, 8.^ However, the mechanism(s) underpinning this are unclear. Given that synaptic events shape brain connectivity^9, 10,^ one possible explanation is that group differences in rsFC patterns are driven by altered neurosignalling. In support of this suggestion, there is evidence of atypicalities in multiple molecular systems in autism^11, 12,^ hinting at a possible causal relationship. Indeed, we have previously shown that several of those systems are likely involved in atypical brain function in autistic individuals^13–28^. Nevertheless, the impact of neurosignalling on a key metric of *large-scale* rsFC in autism has not been directly examined before. Hence, we used our ‘Shiftability’ approach, described in Whelan et al., 2023^29^, to test the hypothesis that *large-scale* rsFC in autism is regulated differently by the µ-opioid system. Specifically, we used a single dose of the atypical antidepressant/anxiolytic tianeptine, a µ-opioid receptor (MOR) agonist^30–32^, to compare autistic and non-autistic brain responses of functional networks with known reproducible differences in autism (across large samples) as reported by Holiga and colleagues^7^.

Tianeptine was selected based on evidence that its µ-opioid receptor target is implicated in autism-relevant processes related to neuronal development and synaptic plasticity^33^ as well as to sensory, reward and social behaviours^12^. Furthermore, the MOR system has been specifically linked to autism by several genetic, case and preclinical studies^12, 34–49.^ We measured *brain-wide* rsFC using the same weighted degree centrality (wDC) index as in Holiga et al.’s^7^ report. wDC is a graph theory measure of node importance in a network. It reflects the sum of all weighted connections between a node and all other nodes^505050^ across the whole brain (1 voxel = 1 node).

We hypothesised that as reported in Holiga et al., 2019^7^, the autism group at baseline would be characterised by hyper-connectivity in higher-order frontoparietal regions and hypo-connectivity in lower-order sensorimotor regions compared to the non-autistic group; however, given our modest sample size in these demanding pharmacological studies, we did not expect a statistically significant difference at baseline. In contrast, as our main focus was on the regulation or ‘responsivity’ of large-scale networks, specifically we anticipated that a) Tianeptine would cause differential wDC shifts in the autistic and non-autistic groups; b) Tianeptine would shift autistic wDC towards baseline non-autistic group levels., i.e. in the autism group, it would decrease wDC of frontoparietal and increase wDC of sensorimotor networks.

## Methods

### Study Ethics and Design

This shiftability^51^ study was conducted in accordance with the Declaration of Helsinki at the Institute of Psychiatry, Psychology and Neuroscience (IoPPN), King’s College London (KCL) in London, UK. Our investigation did not address safety or clinical efficacy and the UK Medicines and Health Regulatory Authority (MHRA) confirmed that the study was not a clinical trial of an Investigational Medicinal Product (IMP) as defined by the EU Directive 2001/20/EC. Nevertheless, the protocol was registered on clinicaltrials.gov for transparency (NCT04145076). Ethical approval was received from a UK Health Research Authority (Stanmore Ethics Committee - 14/LO/0663).

The study evaluated the effects of an acute dose of tianeptine at maximum plasma concentration on rs-fMRI in comparison to inactive placebo in a sample of autistic and non-autistic males.

All participants gave written, informed consent after receiving a complete description of the study. Each person completed two rs-fMRI scan sessions: following administration of either a single 12.5mg dose of encapsulated tianeptine (supplied by Servier Laboratories, Suresnes, France) or a single dose of encapsulated placebo (ascorbic acid), in a randomised, double-blind, crossover design. The randomisation list for the administration order was produced using a computerised random number generator with block randomisation. Scanning commenced approximately 1h after the participant received a dose as tianeptine reaches peak plasma levels 0.94h (±0.47h) following administration^52^. There was a minimum inter-scan interval of eight days to allow for complete drug washout (t_½_ = 2.5h (±1.1h)^52^; washout = 10*t_½_ = 25h). Each participant was examined by a medical doctor prior and subsequent to administration of both doses.

### Participants

Twenty-one non-autistic and 20 autistic males were included in the study. Data from 2 non-autistic participants (2 placebo sessions and 2 drug sessions) and 3 autistic participants (2 placebo sessions and 1 drug session) were excluded from the analysis due to significant head movement, resulting in a final sample of 19 non-autistic and 20 autistic (n_placebo_ = 18; n_drug_ = 19) males.

During the screening phase, participants were excluded if they had a learning disability, any major mental health condition, genetic disorders associated with autism, substance dependence or if they were taking medication targeting the serotonergic systems (at the time of data collection it was still assumed that tianeptine might work as a serotonin-selective reuptake enhancer; evidence for its MOR action was published in 2014^30^ and 2017^32^). Diagnoses of autism spectrum disorder were made by consultant psychiatrists using ICD-10 research criteria^53^ and confirmed using the Autism Diagnostic Interview-Revised (ADI-R)^54^ if an informant was available. Current autistic symptoms were assessed by means of the Autism Diagnostic Observation Schedule (ADOS)^55^. Intelligence quotient (IQ) was measured using the Wechsler Abbreviated Scale of Intelligence test (WASI)^56^.

### MRI Data Collection

MRI data was acquired using a 3T General Electric Discovery MR750 scanner equipped with an 8-channel birdcage head coil at the Centre for Neuroimaging Sciences, IoPPN, KCL, London, UK.

Structural MRI data was collected using a 3D inversion recovery prepared fast spoiled gradient recalled (IR-FSPGR) sequence. Specifically, we used the second-generation sequence^57^ developed by the MRI core of the Alzheimer’s Disease Neuroimaging Initiative (ADNI)^58^ with the following MR parameters: TR = 7312 ms, TE = 3016 ms, TI = 400 ms, FA = 11 °, FoV = 270 mm, matrix size = 256 x 256 voxels, voxel size = 1.055 x 1.055 x 1.2 mm, N of sagittal slices = 196. An echo planar imaging (EPI) sequence with the following MR parameters was used for the acquisition of the resting-state functional MRI data: TR = 2300 ms, TE = 12.7/31/48 ms, FA = 90 °, FoV = 240mm, matrix size = 64 x 64 voxels, voxel size = 3.75 x 3.75 x 4.2 mm, N of axial slices (per volume) = 33, N of volumes = 215, length = 8.24 min.

### MRI Data Pre-processing and Quality Control

The rs-fMRI data was pre-processed using a pipeline nearly identical to the one employed by Holiga et al.,2019^7^. It was implemented in AFNI^59^ (v21.1.07) and consisted of the following steps: despiking, slice time correction, registration of the rs-fMRI image to the structural image, normalisation to standard (i.e. MNI152_T1_2009c) space, tissue segmentation, optimal combination of echoes and motion correction using multi-echo ICA^60, 61,^ smoothing (6mm^3^), scaling (to percent signal change with µ = 0), regression of white matter and cerebrospinal fluid signal, band-pass filter (0.01 - 0.1Hz) and censoring (if framewise displacement > 3mm and/or if motion outlier (i.e. >2 σ)).

The pre-processed data was then passed through a qualitative check using the quality control (QC) reports that are automatically generated by the AFNI software. Specifically, data were inspected for artefact-free structural and functional images, adequate skull-stripping, goodness-of-fit of the registration and the MNI normalisation, presence of defined default mode, visual and auditory networks following seed-based rsFC estimation, sub-threshold percentage of censored volumes (arbitrarily set to 20%), signal coverage similar or superior to the mean coverage.

### Functional Connectivity Estimation and Post-processing

The pre-processed and QC-cleared rs-fMRI data was further processed to obtain a measure of rsFC, namely the graph theory metric used by Holiga et al., 2019^7^ known as weighted degree centrality (wDC). The wDC index represents the sum of weighted connections of each voxel. Accordingly, wDC was calculated for each voxel (in each subject, for each condition) by taking the sum of all correlation coefficients (computed using cosine similarity, S_C_^62, 63^) between the z-score standardised (i.e. µ = 0, σ = 1) time series of a that voxel and the rest of the voxels in the grey matter of the brain (see formula below).

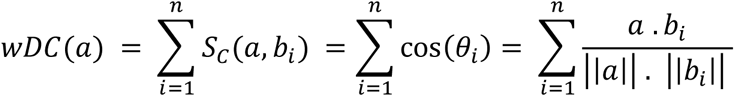

For a given voxel time series vector *a* and the time series vectors of all other voxels *b_i_* in the grey matter of the brain (for a single subject in a single condition), where θ_i_ is the angle between *a* and *b_i_*. **N.B.** *a* and *b_i_* are 205-dimensional vectors as there are 205 time points.

Before estimating wDC, the raw correlation matrices were thresholded (S_C_ > 0.25)^64, 65,^ similar to Holiga et al., 2019^7^, in order to eliminate low temporal correlation as it likely reflects noise.

The resulting wDC images were z-scored, to be referred to as wDC maps. The maps were then constrained to regions that have been previously reported as significantly different between autistic and non-autistic individuals by Holiga et al., 2019^7^. This was achieved by masking out voxels that fall outside of the *EU-AIMS mask* from this 2019 study. The masked wDC maps were further split into two sub-maps: one corresponding to a higher-order network of “hyper-connected” fronto-parietal regions (4446 voxels) and another – to a network of “hypo-connected” sensorimotor regions (2803 voxels). In practice, the *EU-AIMS mask* was first split into its two constituents and each of the sub-masks was resampled to the pre-processed rs-fMRI data, binarised and finally, intersected with the common grey matter mask that was itself generated by intersecting all subject-specific grey matter masks (threshold = 0.85%). The wDC values comprising each sub-mask were averaged to obtain a mean wDC value for each of the two sub-masks in each subject in each condition. The image processing pipeline is summarised in **Figure 1**.

**Figure 1.**
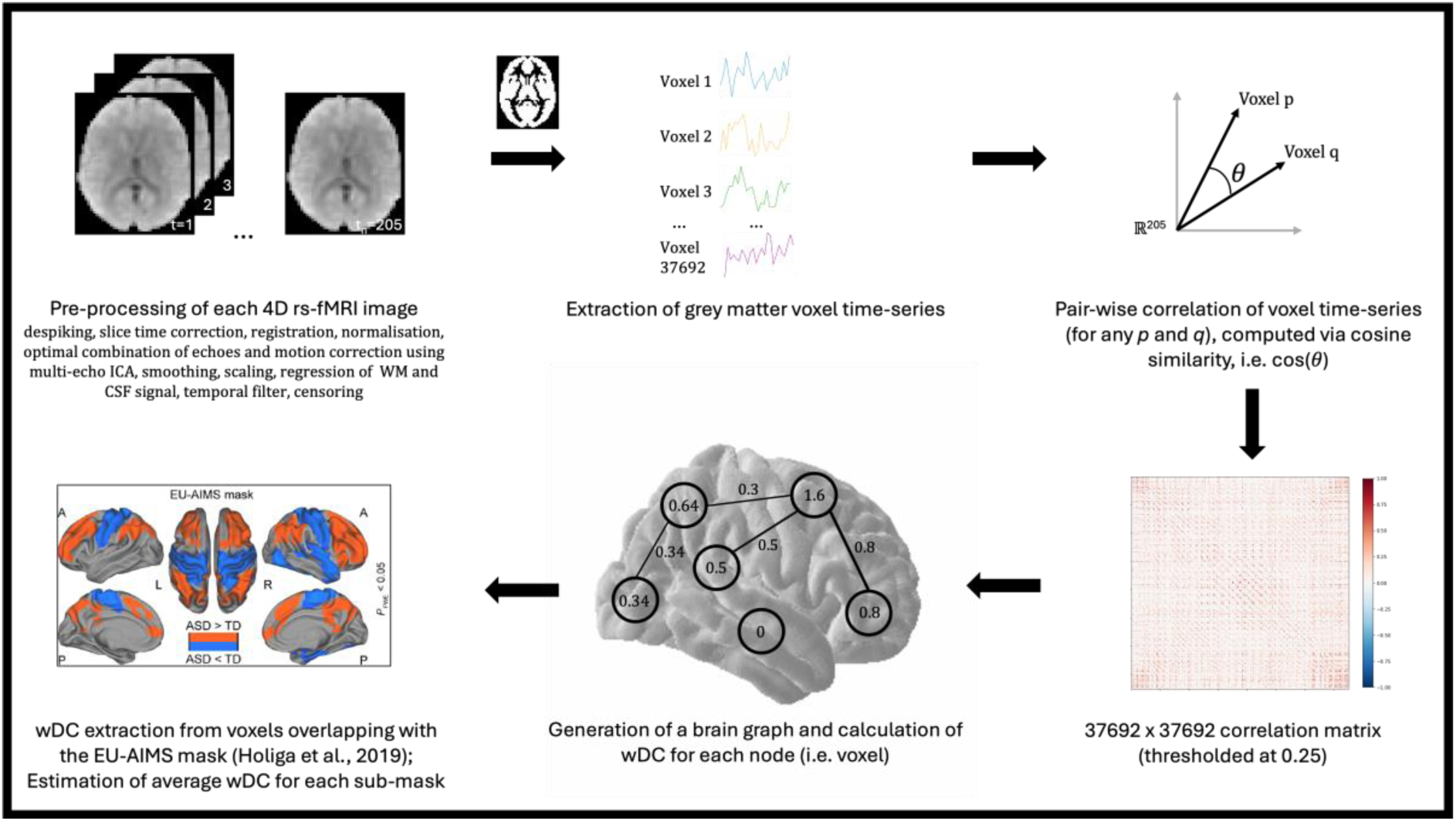
Schematic of the image processing pipeline. EU-AIMS mask image adapted from Holiga et al., 2019^7^ with permission.

### Statistical Analysis

Groups were compared on age, IQ, and in-scanner movement (indexed by mean framewise displacement, mFD). If present, missing data was imputed through mean imputation. Normality was confirmed using a Shapiro-Wilk test in addition to a visual inspection of a frequency distribution histogram and a Q-Q plot. Equality of variances was assessed using either the Bartlett test or the Levene test (for non-normal distributions). Group comparisons were performed using independent or paired two-sample Student t-tests (as appropriate), or in the case of non-normally distributed data – via non-parametric alternatives, i.e. Wilcoxon rank-sum or Wilcoxon signed-rank tests, respectively.

The main effects of group and drug as well as the effect of their interaction on wDC were estimated in a linear mixed effects model (LMM) using restricted maximum likelihood, implemented in R using the *lme4* package. An LMM was fitted for each of the two networks of regions: *y[wDC] ∼ β_0_ + β_1_[group] + β_2_[drug] + β_3_[groupXdrug] + β_4_[mFD] + (1 | Subject ID)*, with *(1 | Subject ID)* representing a random intercept for each subject to take into account within-subject correlation and individual differences in baseline wDC. Another set of LMM’s was fitted to investigate within-group drug effects (2 LMM’s for each of the two networks): *y[wDC] ∼ β_0_ + β_1_[drug] + β_2_[mFD] + (1 | Subject ID)*. Permutation testing was applied to establish the statistical significance of each effect in each model (using *p-testR* in R). Although it has been previously suggested that 1000 permutations are sufficient to produce an accurate approximate permutation test^66^, we performed 5000 permutations of the outcome variable (wDC) to obtain more robust estimates in our relatively small sample. Finally, the outputs of each model were corrected for multiple comparisons using the two-stage Benjamini-Yekutieli FDR method^67^ (R *stats*) to account for potentially complex dependency structures, including negative correlations (α-value = 0.05).

### Code Availability

The rs-fMRI data was pre-processed with AFNI^59^ (v21.1.07).

Computation of weighted degree centrality, data post-processing and assessment of sample characteristics were performed using FSL^68^ commands (only for post-processing) and custom Python v3.7 code, utilising the following auxiliary libraries: *pandas, NumPy, matplotlib, SciPy, Scikit-learn, NiBabel, Nilearn, graph-tool, csv, pickle, os* and *glob*.

Statistical analyses and figure generation were conducted in R using the R Base Package along with the *lme4*, *p-testR, stats*, *ggplot2, MetBrewer*, *tidyr, ggpubr* and *readr* packages. Inkscape (https://inkscape.org) was additionally used to produce the results figure.

All scripts for data processing, analysis and visualisation are available on https://github.com/misho-dimitrov/Tianeptine_wDC.

## Results

### Sample Characteristics

Groups did not differ in terms of age and IQ (**Table 1**). However, the autistic group was characterised by a significantly higher degree of head motion at baseline, as measured by mean framewise displacement, mFD, in mm (**Table 2**). mFD was therefore included as a covariate in each of the linear mixed models assessing µ-opioid-induced shifts in weighted degree centrality (wDC).

**Table 1.** Demographics and clinical characteristics of the sample.

|  | Neurotypical | Autistic | statistic | p-value |
| --- | --- | --- | --- | --- |
| **Demographics** |  |  |  |  |
| **N** | 19 | 20 |  |  |
| **Age [$\mu \pm (SD)$]** | 26 (7) | 29 (10) | w = 150.5 | p = 0.388 |
| **IQ [$\mu \pm (SD)$]** | 114 (10) | 111 (16) | t = 0.659 | p = 0.515 |
| **Clinical Characteristics** |  |  |  |  |
| **ADOS [$\mu \pm (SD)$]** |  |  |  |  |
| Communication | --- | 2 (2) | --- | --- |
| Social Interaction | --- | 6 (3) | --- | --- |
| Imagination | --- | 1 (0) | --- | --- |
| Rep Behaviours | --- | 1 (2) | --- | --- |
| **ADI-R [$\mu \pm (SD)$]** |  |  |  |  |
| Communication | --- | 19 (9) | --- | --- |
| Social Interaction | --- | 13 (6) | --- | --- |
| Rep Behaviours | --- | 5 (2) | --- | --- |

**Table 2.**
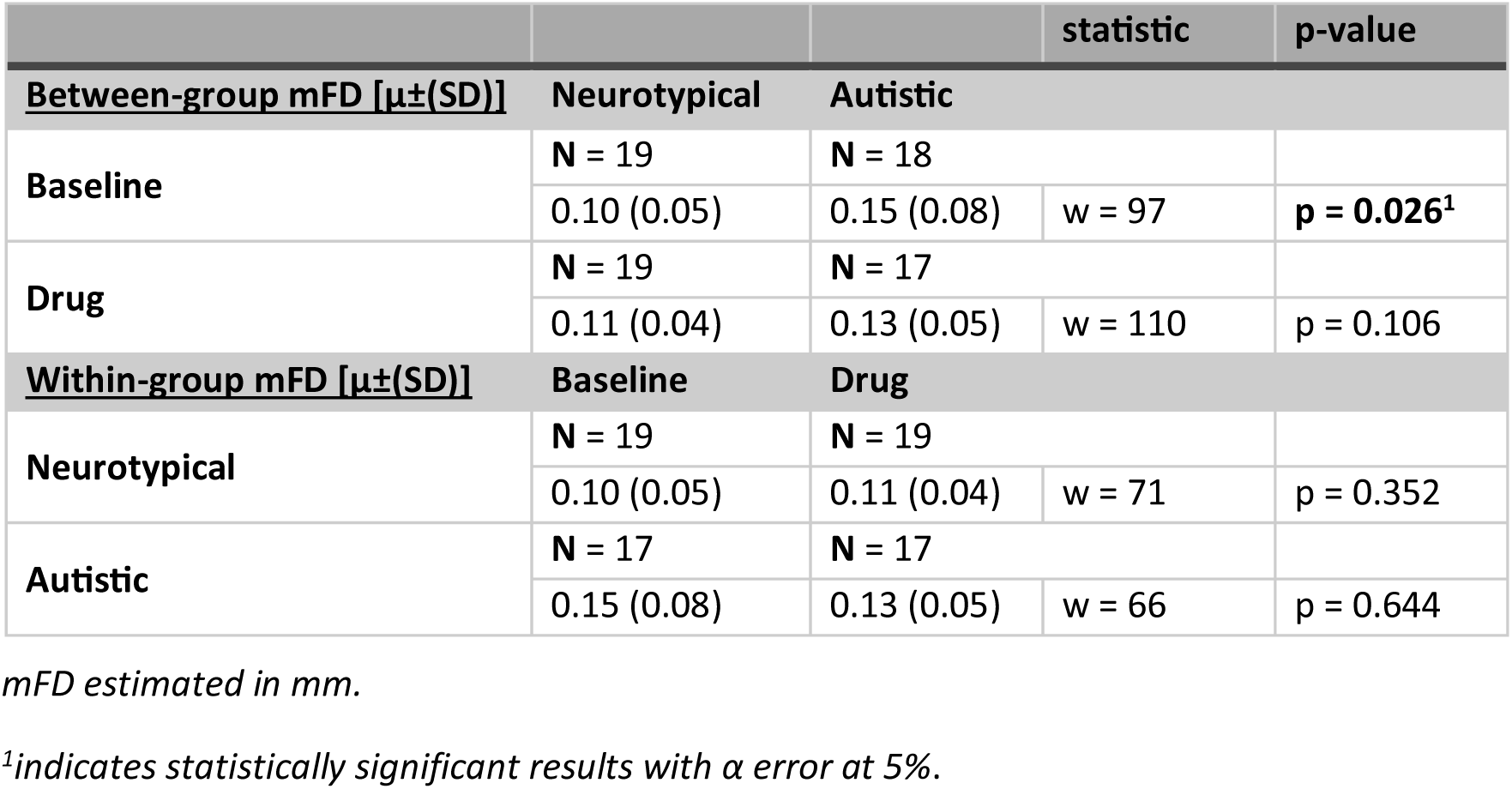
Degree of head motion within the sample.

### Differential µ-opioid Control of Sensorimotor and Higher-Order Networks in Autistic and Non-autistic Individuals

There was no main effect of group for either of the two networks. We observed interaction effects in the hypothesised directions, with a significant sensorimotor shift, which did not, however, survive FDR correction (*t = 2.62; p = 0.185*). The interaction effect on fronto-parietal wDC was not significant (*t = −1.72; p = 0.431*), suggesting that it might be capturing some of the main effects. Consequently, we ran an additional model without the interaction term and compared the model fits. Both the Akaike Information Criterion (−32.78 vs −35.39) and Bayesian Information Criterion (−16.84 vs −21.73) favoured the simpler model. In both instances, the main effect of group was non-significant. Comprehensive linear mixed model output can be found in **Supplementary Table 1**.

### Within-group µ-opioid Shifts

In the non-autistic group, tianeptine did not elicit wDC shifts in fronto-parietal or sensorimotor networks. In both sets of regions, individual trajectories were characterised by a pattern whereby subjects with above-median wDC at baseline tend to undergo positive shifts following drug administration, and vice-versa.

In the autistic group, tianeptine caused a negative wDC shift in fronto-parietal regions, which did not reach statistical significance (*t = −2.18; p = 0.283*). Conversely, the MOR agonist significantly increased sensorimotor wDC (*t = 3.72; **p = 0.043***). The same pattern that was present in the individual trajectories of the non-autistic subjects was also noted in the autistic group, although it was limited to fronto-parietal areas. In the sensorimotor network, we observed a general increase in wDC, regardless of baseline wDC.

Visual representation of the results is shown in **Figure 2**. Detailed output from the linear mixed models can be found in **Supplementary Tables 2** and **3**.

**Figure 2.**
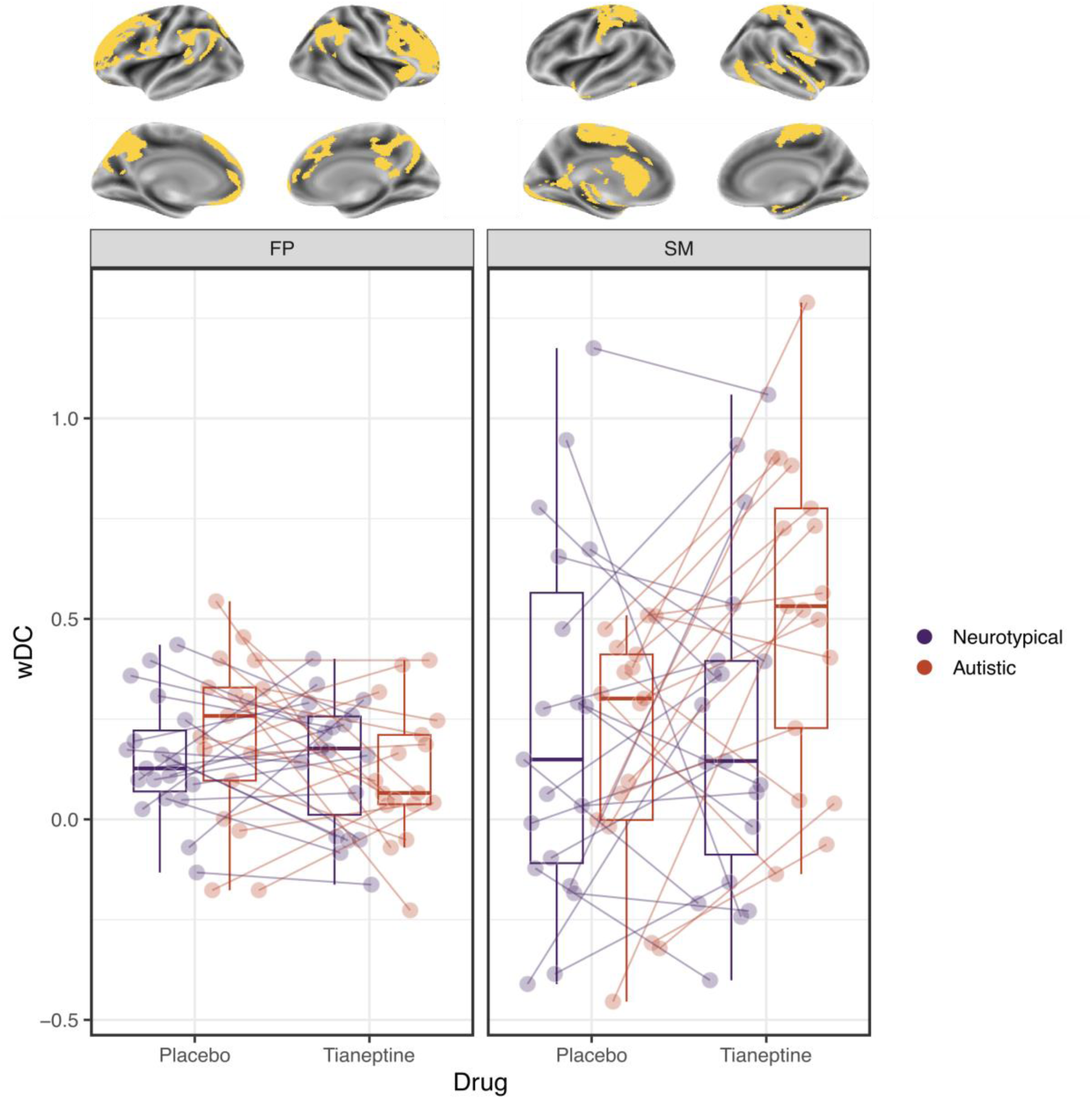
The effect of tianeptine on wDC averaged across fronto-parietal (left) and sensorimotor (right) brain regions in neurotypical and autistic individuals. The brain plots (top) are showing the regions comprising each mask. wDC – weighted degree centrality; FP – fronto-parietal; SM - sensorimotor.

## Discussion

We found that in the autistic group, administration of the µ-opioid receptor (MOR) agonist tianeptine shifted sensorimotor weighted degree centrality (wDC), a network measure of node importance. This particular cluster of sensorimotor regions has been previously reported as atypically hypo-connected (indexed by decreased wDC) by Holiga and colleagues using data from several large-scale studies comparing autistic to non-autistic individuals in a wide age range^7^. Here, we showed that MOR modulation can *increase* wDC in this specific cluster. This suggests that reproducible functional brain atypicalities in sensorimotor regions of the autistic brain observed in large-scale studies might be driven by atypical MOR function.

Differences in sensorimotor processing and behaviours are commonly reported in the autistic population^69–71^ and form part of the diagnostic criteria for Autism Spectrum Disorder in the DSM-5^72^ and ICD-11^73^. Unusual sensorimotor features are amongst the earliest signs of autism^74–76^, persist throughout life^72^ and exert possible cascading effects on higher-order social and cognitive characteristics of this neurodevelopmental condition^69–71, 77.^ In addition, the presence of sensory atypicalities in autistic individuals can negatively affect adaptive behaviour^78, 79^ and has been linked to mental health conditions such as depression^80, 81^ and anxiety^82–84^, further highlighting the practical importance of elucidating sensorimotor mechanisms in autism.

These behavioural and clinical observations are mirrored in functional brain differences as well. For instance, there is evidence pointing to atypical sensorimotor resting-state functional connectivity (rsFC), reflected by an altered *Regional Homogeneity* index, in the brains of neonates with an increased likelihood of autism diagnosis^85^. Preterm birth, which has been linked to a greater chance of atypical neurodevelopment^86^ including autism^87–92^, has also been associated with an atypical dynamic rsFC profile of a sensorimotor network in the neonatal brain that are predictive of neurodevelopmental and autistic traits at 18 months^93^. This atypical sensorimotor rsFC feature appears to be present in the autistic population across a wide age range, as shown by recent large-scale rsFC studies^7, 8.^ Furthermore, it has been reported as the most informative predictor in machine learning-based diagnostic classification^94^ and has been linked to differences in sensory processing, social difficulties, and restricted and repetitive behaviours^8^. These strands of evidence lend support to the idea that sensorimotor atypicalities in autism can be observed on a behavioural *as well as* on a biological level, whereby the latter can not only precede but also potentially predict and explain the former.

What remains unclear, however, is how these differences in behaviour and underlying gross brain function of sensorimotor systems are themselves underpinned by neurosignalling at a more fundamental level. An underexplored neurosignalling candidate in autism research is the µ-opioid receptor (MOR) system, as previous studies have mainly focused on GABA, glutamate and serotonin (5-HT), for instance^11^. MOR plays a role in several inter-connected processes, including sensory processing, social behaviour and reward, all of which are known to differ in autism^12^. Emerging evidence suggests that the MOR system likely functions atypically in autism^12, 34–49^, making it a promising target for investigation. Mechanistically, the µ-opioid receptor is a also plausible molecular modulator of the functional landscape of sensorimotor networks in the brain given its moderate availability in those regions (at least in non-autistic individuals)^95, 96^. Combined with our results showing that the MOR agonist tianeptine can increase sensorimotor wDC (previously shown to be decreased in autistic individuals^7^), this suggests that the MOR system might contribute to characteristics and behaviours associated with autism. Notably, even if this influence is confined to sensorimotor regions and in turn, sensorimotor behaviours, knock-on effects on other systems known to be atypical in autism might also be present. These effects might contribute to social and cognitive differences and potentially predispose individuals to conditions such as depression and anxiety, which are also associated with MOR atypicalities^97^. As such, while preliminary, these findings possess a potential translational value and thus, warrant further investigation.

Nevertheless, although there are several lines of evidence that emphasize tianeptine MOR agonist actions^30–32^, there are also reports of various other direct (∂-opioid)^30^ and indirect effects on other systems, including GABA^31^, glutamate^98–101^, dopamine^31, 102, 103^, serotonin^104–110^ and BDNF-TrkB^111, 112^. Each of these pathways has been found to be atypical in autism^11, 113,^ raising the question of whether the shifts that we observed in this study can be solely attributed to direct MOR effects or whether they can be a consequence of a broader multi-system action. It is possible that the two are inseparable given well-documented interactions between MOR and GABA, glutamate, dopamine, serotonin^114^, BDNF-TrkB^115^.

This brings us to another consideration: to date, the majority of drugs that have been tested in autistic individuals to address core and/or co-occurring difficulties have primarily targeted single neurosignalling systems^116^. However, autism is a complex condition linked to atypicalities across multiple neural systems^11^ as well as in the overall excitation/inhibition balance^117, 118^ in the brain. Therefore, it is possible that the current dearth of effective and targeted pharmacological approaches in the context of autism^5, 116^ may partly stem from a mismatch between the complexity of behavioural phenotypes and compounds targeting them.

Despite the putative multi-pathway effects of tianeptine, its primary action is most likely via the µ-opioid receptor, providing a strong indication that this receptor system might be uniquely positioned to modulate sensorimotor functionality in the human brain, with possible practical implications for autism specifically. Further research is needed to corroborate this and establish whether it has any translational value.

### Limitations

Our findings should be interpreted alongside several key limitations. The analysis was based on a modest sample size dictated by stringent inclusion and exclusion criteria as well as stringent motion correction criteria. However, the repeated measures design of the study improved power by decreasing the heterogeneity of the sample. Nevertheless, larger samples might provide more accurate estimates of tianeptine’s effects on the human brain. Our investigation was limited to adult males. While this provides an opportunity to probe neurosignalling mechanisms in a more homogenous sample, it reduces the generalisability of our findings. This has several possible implications. For instance, although the adult brain is characterised by a certain level of plasticity, it is nonetheless a system with established homeostatic properties^119^. As such, it might be less susceptible to external influences and thus, might respond differently to pharmacological perturbations compared to the child or adolescent brain. Additionally, previous research has highlighted sex differences in brain structure, function and connectivity in the autistic population^120–124^ as well as sex differences in pharmacological responses in neurotypical individuals^125^. This implies that results obtained in autistic males may not directly translate to autistic females. Hence, it would be important for future studies to extend our work to include other sub-groups of the autistic population. Finally, this study employed an acute 12.5mg dose of tianeptine. However, a single dose may not produce effects comparable to those of a sustained administration regimen (e.g. the standard 12.5mg TID used in clinical practice), be it in terms of extent, magnitude or direction. In addition, evidence suggests that the levels of certain molecular targets of tianeptine may vary seasonally^126, 127,^ which could introduce potential bias in a cross-sectional drug study design like ours. Further research is needed to address these unresolved questions.

## Conclusions

In this study, we demonstrated that the atypical µ-opioid agonist tianeptine increases the centrality of sensorimotor regions previously identified as having *reduced* centrality in autism at baseline. Given the impact of sensorimotor atypicalities on other core features of autism, adaptability, and mental health, our findings highlight the potential relevance of the µ-opioid system for further investigation. While our results suggest that µ-opioid neurosignalling may underlie atypical sensorimotor function in autism, whether these findings have practical implications for developing more targeted interventions remain an open question for future translational research.

## Data Availability

Data from this study is available upon request.

## Author Contributions

**MD**, **JLF**, **RHW**, **DGMM**, **GMM** and **ED** designed the study. **JLF** and **RHW** collected the data with support from **VS**, **DMR**, **SB** and **GI**. **NMLW**, **SL**, **LGSF**, **IV**, **JS**, **DJL**, **SH** and **DB** provided technical support. **MD** processed and analysed the data. **MD**, **SH**, **MDT**, **DB**, **DGMM**, **GMM** and **ED** interpreted the data. **MD** drafted the manuscript in consultation with **DGMM**, **GMM** and **ED**.

All authors read and provided critical feedback on the manuscript, and approved the final version.

