## Supplementary Information for "µ-Opioid Modulation of Sensorimotor Functional Connectivity in Autism: Insights from a Pharmacological Neuroimaging Investigation using Tianeptine"

Supplementary materials

**Supplementary Table 1.** Results from the interaction analysis. FDR correction has only been applied to effects of interest. Statistically significant effects are highlighted in bold.

| **term** | **estimate** | **t-statistic** | **p-value (perm)** | **p-value (FDR)** |
| --- | --- | --- | --- | --- |
| **Fronto-parietal** |  |  |  |  |
| (Intercept) | 0.27 | 5.35 | 0.006 | **-** |
| drug | <0.01 | 0.08 | 0.937 | - |
| group | 0.11 | 2.04 | 0.043 | 0.283 |
| group X drug | -0.12 | -1.72 | 0.099 | 0.431 |
| mFD | -1.17 | -3.43 | **<0.001** | **-** |
| **Sensorimotor** |  |  |  |  |
| (Intercept) | 0.06 | 0.52 | 0.972 | - |
| drug | -0.01 | -0.14 | 0.895 | - |
| group | -0.13 | -0.98 | 0.335 | 1.000 |
| group X drug | 0.38 | 2.62 | **0.017** | 0.185 |
| mFD | 1.69 | 2.03 | **0.042** | **-** |

**Supplementary Table 2.** The effect of tianeptine on wDC in the non-autistic group. FDR correction has only been applied to effects of interest.

| **term** | **estimate** | **t-statistic** | **p-value (perm)** | **p-value (FDR)** |
| --- | --- | --- | --- | --- |
| **Fronto-parietal** |  |  |  |  |
| (Intercept) | 0.21 | 2.86 | 0.243 | - |
| drug | <0.01 | 0.03 | 0.980 | 1.000 |
| mFD | -0.55 | -0.89 | 0.361 | - |
| **Sensorimotor** |  |  |  |  |
| (Intercept) | -0.01 | -0.05 | 0.993 | - |
| drug | -0.02 | -0.16 | 0.872 | 1.000 |
| mFD | 2.39 | 1.42 | 0.150 | - |

**Supplementary Table 3**. The effect of tianeptine on wDC in the autistic group. FDR correction has only been applied to effects of interest. Statistically significant effects are highlighted in bold.

| **term** | **estimate** | **t-statistic** | **p-value (perm)** | **p-value (FDR)** |
| --- | --- | --- | --- | --- |
| **Fronto-parietal** |  |  |  |  |
| (Intercept) | 0.43 | 6.06 | 0.000 | - |
| drug | -0.12 | -2.18 | 0.052 | 0.283 |
| mFD | -1.51 | -3.70 | **0.001** | - |
| **Sensorimotor** |  |  |  |  |
| (Intercept) | -0.03 | -0.17 | 0.989 | - |
| drug | 0.36 | 3.72 | **0.002** | **0.043** |
| mFD | 1.39 | 1.57 | 0.148 | - |
